# Mental Disorder Prevalence Among Populations Impacted by Coronavirus Pandemics: A Multilevel Meta-Analytic Study of COVID-19, MERS & SARS

**DOI:** 10.1101/2020.12.18.20248499

**Authors:** Matt Boden, Nichole Cohen, Jessilyn M. Froelich, Katherine J. Hoggatt, Hoda S. Abdel Magid, Swapandeep S. Mushiana

## Abstract

**Objective:** Through a systematic review and meta-analysis of research on COVID-19, severe acute respiratory syndrome (SARS) and middle east respiratory syndrome (MERS) pandemics, we investigated whether mental disorder prevalence: (a) was elevated among populations impacted by coronavirus pandemics (relative to unselected populations reported in the literature), and (b) varied by disorder (undifferentiated psychiatric morbidity, anxiety, depressive, posttraumatic stress disorders [PTSD]) and impacted population (community, infected/recovered, healthcare provider, quarantined).

**Method:** From 68 publications (*N*=87,586 participants), 808 estimates were included in a series of multilevel meta-analyses/regressions including random effects to account for estimates nested within studies.

**Results:** Median summary point prevalence estimates varied by disorder and population. Psychiatric morbidity (20%-56%), PTSD (10-26%) and depression (9-27%) were most prevalent in most populations. The highest prevalence of each disorder was found among infected/recovered adults (18-56%), followed by healthcare providers (11-28%) and community adults (11-20%). Prevalence estimates were often notably higher than reported for unselected samples. Sensitivity analyses demonstrated that overall prevalence estimates moderately varied by pandemic, study location, and mental disorder measure type.

**Conclusion:** Coronavirus pandemics are associated with multiple mental disorders in several impacted populations. Needed are investigations of causal links between specific pandemic-related stressors, threats, and traumas and mental disorders.

## 1. Introduction

Numerous studies document the often-substantial adverse mental health impact of coronavirus pandemics (Brooks et al., 2020; Mak et al., 2009). With the onset of COVID-19, useful information will be gained from a systematic review of this literature and quantitative summary of prevalence of mental disorders that may be common to populations impacted by coronaviruses. Through a systematic review and meta-analysis of COVID-19, severe acute respiratory syndrome (SARS) and middle east respiratory syndrome (MERS) pandemic research literatures, we addressed two questions. First, is mental disorder prevalence elevated among populations impacted by coronavirus pandemics relative to unselected populations reported in the literature? Second, does mental disorder prevalence vary by disorder (undifferentiated psychiatric morbidity, anxiety, depressive, posttraumatic stress disorders [PTSD]) and impacted population (community, infected/recovered, healthcare provider, quarantined)? We focused our investigation on disorders that are common in the general population and/or theoretically or empirically linked to stressors, threats and traumas that we hypothesized would be prevalent during coronavirus pandemics (see below). As the mental health impact of a coronavirus might not manifest in distinguishable disorders in the acute phases of pandemics, we also investigated undifferentiated psychiatric morbidity (i.e., distress commensurate with a mental disorder diagnosis).

Physical exposure to a virus (e.g., through job duties), media exposure, exposure to illness and death, movement restrictions, interpersonal loss, and (for COVID-19) unemployment and economic deprivation are key pandemic-related threats, stressors and traumas that are likely to increase the risk of a mental disorder (Brooks et al., 2020; Dooley et al., 1994; Garfin et al., 2020; Lai et al., 2020; Liu et al., 2012). Populations experiencing a greater frequency or intensity of pandemic-related stressors are likely to be at greater risk of experiencing a mental disorder (Galea et al., 2020). Among adults infected with a coronavirus, threats to health and mortality, and disruption to routines (e.g., absence from work) will be pronounced and more impactful the greater the severity of infection (Bienvenu et al., 2018; Wu et al., 2005). Functional impairment and disability may further increase risk for mental disorders among recovered adults (Lam et al., 2009; Lancee et al., 2008; Lee et al., 2007; Lee et al., 2019; Mak et al., 2009). Healthcare providers, especially front-line treatment providers who contend with threats, stressors and traumas such as repeated exposure to infected and dying people and morally ambiguous decisions regarding who receives treatment (Lai et al., 2020; Lancee et al., 2008; Liu et al., 2012) may be at increased risk of mental disorders. Quarantined adults may be at increased risk of mental disorders due to threats to health, lack of social contact, and disruptions to routine (Brooks et al., 2020). Anxiety and depressive disorders are likely to be common as any given person may experience multiple threats and stressors that contribute to such disorders (Cisler et al., 2010; Hammen, 2005). Infected/recovered adults and healthcare providers, in particular, may experience traumatic events (e.g., invasive treatments, witnessing death) that increase risk of post-traumatic stress disorder (PTSD; Lai et al., 2020; Mak et al., 2009).

Based on a systematic review of COVID-19, SARS, and MERS research, we conducted multilevel meta-analyses/regressions to derive summary prevalence estimates for multiple disorders (undifferentiated psychiatric morbidity, anxiety, depressive, PTSD) in adult populations (community [including students], infected/recovered, healthcare provider, quarantined). We investigated whether mental disorder prevalence was elevated among populations impacted by coronavirus pandemics relative to unselected populations reported in the literature. For example, a meta-analysis of 157 studies from 59 countries (*N ∼*660,000; Steel et al., 2014) found a 12-month prevalence of 15.4% for combined mood (including bipolar) and anxiety (including PTSD) disorders. Twelve-month (or less) prevalence has also been reported in epidemiological studies including unselected samples representative of the adult populations of: (a) China (any disorder=9.3; anxiety disorder=5.0; depressive disorder=3.6; PTSD=.2%; Huang et al., 2019), (b) Europe (anxiety disorder=6.4, major depression=3.9; ESEMeD/MHEDEA 2000 Investigators, 2004), and (c) the United States (any disorder=26.2, anxiety disorder=18.1, depressive disorder=6.7, PTSD=3.5; Kessler et al., 2005). We hypothesized that, relative to these estimates, all disorders would be more prevalent in all populations that we investigated given the frequent and often impactful threats, stressors and traumas associated with coronavirus pandemics. As the frequency and impact of particular threats, stressors and traumas is likely to vary in type and by population, we further hypothesized that mental disorder prevalence would vary by disorder and impacted population.

As individual studies often provided estimates of multiple disorders from the same population, we utilized three-level meta-analytic models and included random effects to account for estimates nested within studies. The use of multilevel models allowed us to include all relevant prevalence estimates (e.g., by sex, age, income-level), thus further maximizing the information provided by any given study. By investigating mental disorder prevalence in multiple populations potentially impacted by coronavirus pandemics, our study provides information not found in published meta-analyses related to the aims of the current paper (de Pablo et al., 2020; Rogers et al., 2020), which focused on single populations (healthcare providers, severely infected/recovered). Comparison of mental disorder prevalence across populations can inform targeted research on populations most impacted by coronavirus, and downstream resource allocation to populations most in need.

## 2. Method

This study is part of a broad registered protocol (Boden, 2020), and was conducted according to PRISMA (Moher et al., 2009) and MOOSE guidelines (Stroup et al, 2000).

### 2.1. Literature Search Strategy & Criteria

From April 15, 2020 until June 1, 2020, two study staff (MB, NC) conducted a key-word search of electronic databases PubMED, PsychINFO and Google Scholar (with focused searches by author additionally conducted in Scopus and Web of Science) for peer-reviewed, English language publications. We searched for a broad set of studies by forming all combinations of key words (a) avian flu, coronavirus, COVID, Ebola, equine flu, flu, H1N1, Influenza, MERS, quarantine, swine flu, SARS, respiratory, crossed with (b) emotional distress, mental health, anxiety, depression, psychological distress, posttraumatic (and separately, post traumatic and PTSD/PTS/PTST, the latter on the advice of a peer-reviewer). Because the literature search was intended to inform several projects in addition to this meta-analysis, search terms unrelated to coronavirus pandemics were included (e.g., Ebola; Boden, 2020). We examined reference lists of identified publications and lists of cited studies to identify additional studies.

### 2.2. Study Sample

Studies meeting the following criteria were included in this meta-analysis: (1) available in English language, (2) peer-reviewed (no grey literature or preprints), (3) reported source data (no reviews), (4) focused on mental disorders (undifferentiated psychiatric morbidity, anxiety, depression or PTSD) related to COVID-19, MERS, SARS, (5) included quantitative estimates of mental disorder prevalence assessed by clinician diagnosis or measures with published psychometric validation data, (6) included adult participants (age >=18). Electronic database searches yielded a total of 4,142 publications that potentially met inclusion criteria. Screening of titles and abstracts yielded 279 studies that were coded (including studies of non-coronavirus outbreaks/epidemics coded as part of the broader project), with 68 studies meeting inclusion criteria (see Figure 1).

**Figure 1.**
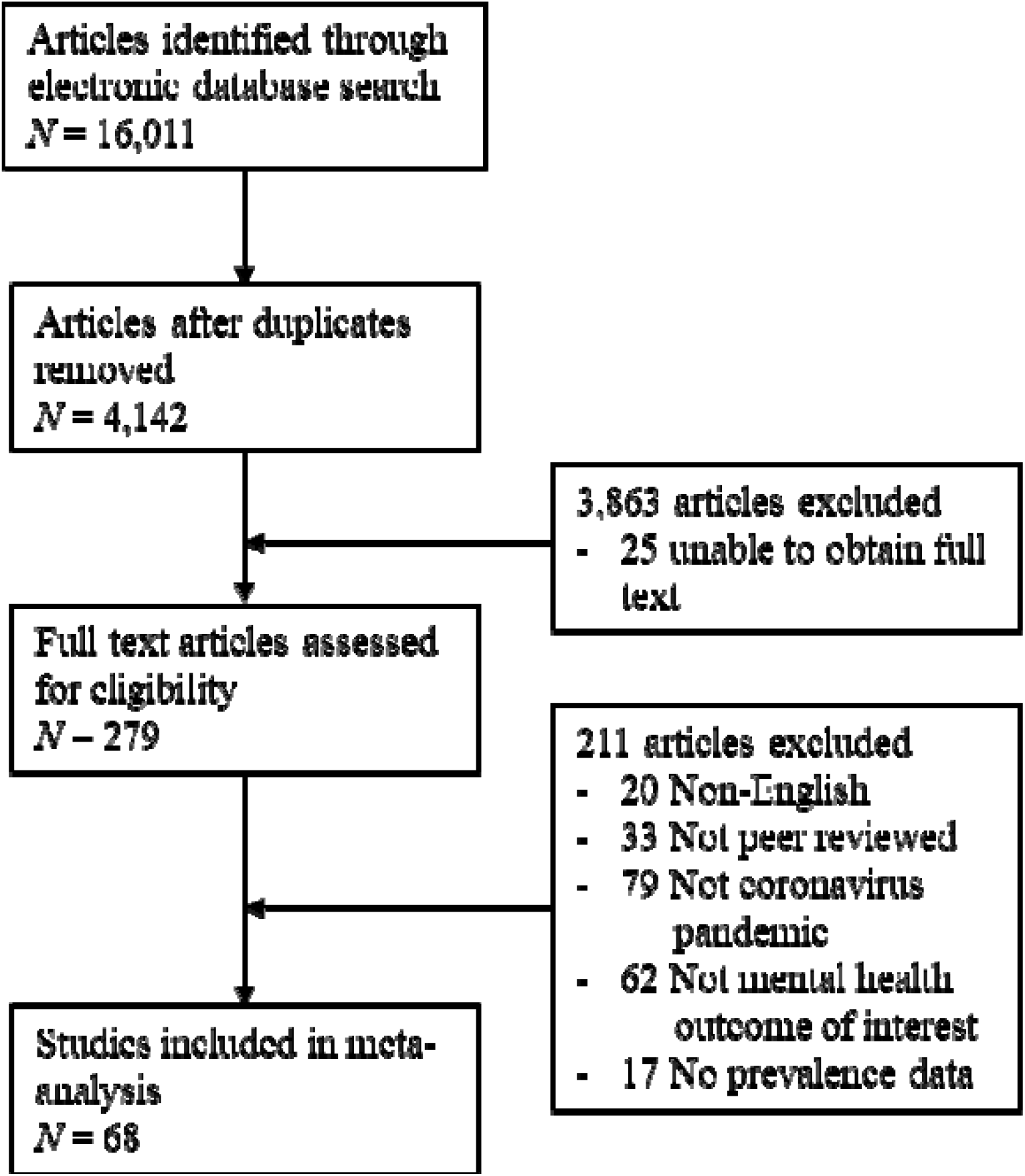
Study screening, selection, coding and analysis.

### 2.3. Coding of Study and Effect Characteristics

Publications were first coded by one of four study authors (MB, NC, SM, JF), with the majority coded by the first author. The first author developed the coding scheme and trained all other study coders through didactics, iterative feedback on coding of a subset of studies, and confirmation of codes by the first author. Study coders identified studies that met inclusion criteria, and extracted data based on the coding scheme developed the first author. A subset of studies (144/279) was coded a second time to ensure consistency of coding. Disagreements between primary and secondary codes were resolved by the first author.

Coded study attributes include mental disorder prevalence in terms of: (a) sample size and (b) size of sample positive for a disorder, (c) type of disorder (undifferentiated psychiatric morbidity, anxiety, depression, PTSD), (d) sample population (community [including university students], infected/recovered, healthcare providers, quarantined), (e) pandemic (COVID-19, SARS, MERS), (f) location (Canada, China, Hong Kong, Korea, Saudi Arabia, Singapore, Taiwan, Italy, United Stated, Multi: Singapore and India), (g) timing (acute, within 1-year of main outbreak, more than 1-year after main outbreak), and (h) measures (see Supplement A). Measures of psychiatric morbidity included questionnaires assessing psychological/emotional distress and multiple types of psychopathology (e.g., Hospital Anxiety and Depression Scale) and clinician assessment of an unspecified mental disorder.

We coded all relevant estimates provided in each study, including estimates for different disorders and the same disorder assessed by different measures, at different time-points, or among different populations and subpopulations (e.g., quarantined versus community, males versus females, doctors versus nurses). Thus, we coded sociodemographic and socioeconomic characteristics but did not include them in moderator analyses due to inconsistent reporting across studies. We also coded study attributes not included in the reported analyses (i.e., means and standard deviation for continuous measures of mental disorders/symptoms).

We manually calculated size of sample positive for a disorder when not reported by the study. For questionnaire measures, we coded size of sample positive for a disorder consistent with the study authors, who typically utilized established cutoffs reported in prior research. Estimates obtained from non-standard cutoffs were identified for sensitivity analysis. When severity thresholds (only) were provided (e.g., mild, moderate, severe), participants in moderate or higher categories were coded as positive for a disorder unless otherwise specified in prior research. Estimates obtained from total but not subscale scores (e.g., PTSD hyperarousal) were included in analyses.

Study quality was assessed by the Systematic Assessment of Quality in Observation Research (SAQOR) tool (Ross et al., 2011) with several changes to increase the applicability of the system to our coding of prevalence (see Supplement B).

### 2.4. Computation of Effect Sizes and Data Analysis

Using the metafor (Viechtbauer, 2010) package in R, we conducted a series of mixed-effect multilevel meta-analyses/regressions. Our main analyses consisted of four meta-analyses/regressions, each of which included a random effect for studies and estimates within studies. First, we conducted a meta-analysis to obtain an overall summary prevalence estimate across disorders and populations. Second, we conducted a meta-regression including (dummy-coded) *disorder* as a fixed effect moderator to obtain summary prevalence of each disorder across populations. We conducted a third meta-regression including (dummy-coded) *population* to obtain overall mental disorder prevalence in each population. The fourth meta-regression, including dummy-coded *disorder, population*, and their product, provided summary estimates of individual disorders in individual populations. Studies that utilized the same samples were analyzed as a single study.

Models were implemented using restricted maximum-likelihood estimation. Study prevalence estimates (i.e., sample size with disorder divided by total sample size) and 95% confidence intervals (CI) were Freeman-Tukey double arcsine transformed (Freeman et al., 1950) for improved statistical properties. Fixed effects and CI were back-transformed (Miller, 1978) to provide summary prevalence estimates. The *I*^2^ statistic measured percentage of heterogeneity due to true between-study and between-estimate differences, and the interclass correlation coefficient (ICC), the association between underlying estimates. Profile plots indicated whether all variance components were statistically identifiable.

As part of sensitivity/quality analyses, we re-conducted our four main analyses using a cluster-robust estimator of standard errors (similar to the Eicker-Huber-White method) useful when multiple, dependent outcomes come from individual studies (Hedges et al., 2010). Additionally, in a series of meta-regressions, we examined whether the overall summary effect varied by: (1) pandemic, (2) location, (3) timing, (4) measure (self-report questionnaire versus clinician assessment; for a related example see Anderson et al., 2001), (5) scoring (non-standard/unknown versus standard scoring of measure), and (6) study quality (very low, low, moderate). Analysis of publication bias was not conducted as most studies did not use inferential statistics to test hypotheses regarding prevalence, and thus, were not more likely to be published because of statistically significant results.

## 3. Results

From 68 publications (of which six studies/three-pairs utilized the same or partially overlapping samples) including 87,586 participants, 808 individual estimates were obtained. Summing *I*^*2*^ values representing between- and within-study heterogeneity, approximately 99% of variance between- and within-studies was due to true variation rather than sampling error in main analyses. As expected, between-study heterogeneity was larger (Range *I*^*2*^ = 72.7-73.6%) than within-study heterogeneity (Range *I*^*2*^ = 25.0-26.2%). ICC ranged from .74 to .75, indicating a strong association between estimates within studies. Models with random effects fit significantly better than respective models without (all *p*<.001). Profile plots indicated all variance components were statistically identifiable. Summary estimates for individual disorders/populations were based on varying numbers of studies, individual effects and participants (see Table 1). CI were large for summary estimates derived from few studies/estimates (e.g., quarantined), thus indicating greater imprecision.

**Table 1.** Sample size and Meta-Analysis/Regression Results, Including Summary Estimates/Prevalence and 95% Confidence Intervals by Disorder and Population.

| Population | Disorder | Study *N* | IE *N* | Sample *N* | SE | 95% CI<br>LB | 95% CI<br>UB | SPP | SPP 95%<br>CI LB | SPP 95%<br>CI UB |
| --- | --- | --- | --- | --- | --- | --- | --- | --- | --- | --- |
| All | All | 65* | 808 | 87586 | .46 | .41 | .51 | .19 | .15 | .23 |
|  | Psych. morbidity | 20 | 232 | 10818 | .60 | .55 | .65 | .31 | .26 | .36 |
|  | Anxiety | 29 | 168 | 33263 | .37 | .32 | .41 | .12 | .09 | .15 |
|  | Depression | 31 | 162 | 24794 | .44 | .39 | .49 | .17 | .14 | .21 |
|  | PTSD | 39 | 246 | 18711 | .45 | .40 | .49 | .18 | .14 | .22 |
| Community | All | 21 | 210 | 49229 | .42 | .35 | .49 | .16 | .11 | .21 |
|  | Psych. morbidity | 4 | 35 | 4446 | .47 | .38 | .57 | .20 | .13 | .28 |
|  | Anxiety | 11 | 85 | 21538 | .39 | .32 | .46 | .13 | .09 | .19 |
|  | Depression | 10 | 46 | 13685 | .44 | .36 | .51 | .17 | .12 | .23 |
|  | PTSD | 11 | 44 | 9560 | .36 | .28 | .43 | .11 | .07 | .17 |
| Infected/recovered | All | 25 | 352 | 30191 | .57 | .48 | .66 | .29 | .21 | .38 |
|  | Psych. morbidity | 11 | 108 | 5664 | .84 | .74 | .95 | .56 | .46 | .66 |
|  | Anxiety | 8 | 40 | 8537 | .45 | .36 | .54 | .18 | .11 | .26 |
|  | Depression | 9 | 79 | 9399 | .56 | .47 | .65 | .27 | .19 | .36 |
|  | PTSD | 16 | 125 | 6591 | .55 | .46 | .64 | .26 | .19 | .35 |
| Healthcare providers | All | 14 | 173 | 3375 | .46 | .39 | .52 | .19 | .14 | .24 |
|  | Psych. morbidity | 4 | 31 | 521 | .56 | .49 | .63 | .28 | .22 | .34 |
|  | Anxiety | 8 | 38 | 944 | .35 | .28 | .42 | .11 | .06 | .16 |
|  | Depression | 9 | 33 | 1000 | .42 | .36 | .49 | .16 | .11 | .21 |
|  | PTSD | 9 | 71 | 910 | .47 | .40 | .53 | .20 | .15 | .25 |
| Quarantined | All | 6 | 73 | 4791 | .34 | .22 | .46 | .10 | .04 | .19 |
|  | Psych. morbidity | 1 | 58 | 187 | .56 | .24 | .88 | .28 | .05 | .60 |
|  | Anxiety | 3 | 5 | 2244 | .22 | .08 | .37 | .04 | .00 | .12 |
|  | Depression | 3 | 4 | 710 | .33 | .16 | .49 | .09 | .02 | .21 |
|  | PTSD | 3 | 6 | 1650 | .34 | .19 | .48 | .10 | .03 | .21 |

\* Independent studies reporting data from the same sample ( $N=3$ pairs) were coded as single studies that provided multiple effect sizes. IE = Individual estimate. SE = Summary estimate. CI = Confidence interval. LB = Lower bound. UB = Upper bound. SPP = Summary point prevalence (i.e., back-transformed Freeman-Tukey double arcsine transformed estimates; Miller, 1975).

The median summary point prevalence for mental disorders across disorders and populations was 19% (95%CI=15-23%). Prevalence estimates of 20% or higher were found for 9 (36%) disorders/populations with a high of 56% for psychiatric morbidity among infected/recovered adults (see Table 1). Occurring in 31% (95%CI=26-36%) of the overall sample, psychiatric morbidity had the highest prevalence across all populations (see Figure 2). Across populations, PTSD had the second highest prevalence (18%, 95%CI=14-22%), closely followed by depression (17%, 95%CI=14-21%) and anxiety (12%, 95%CI=9-15%).

**Figure 2.**
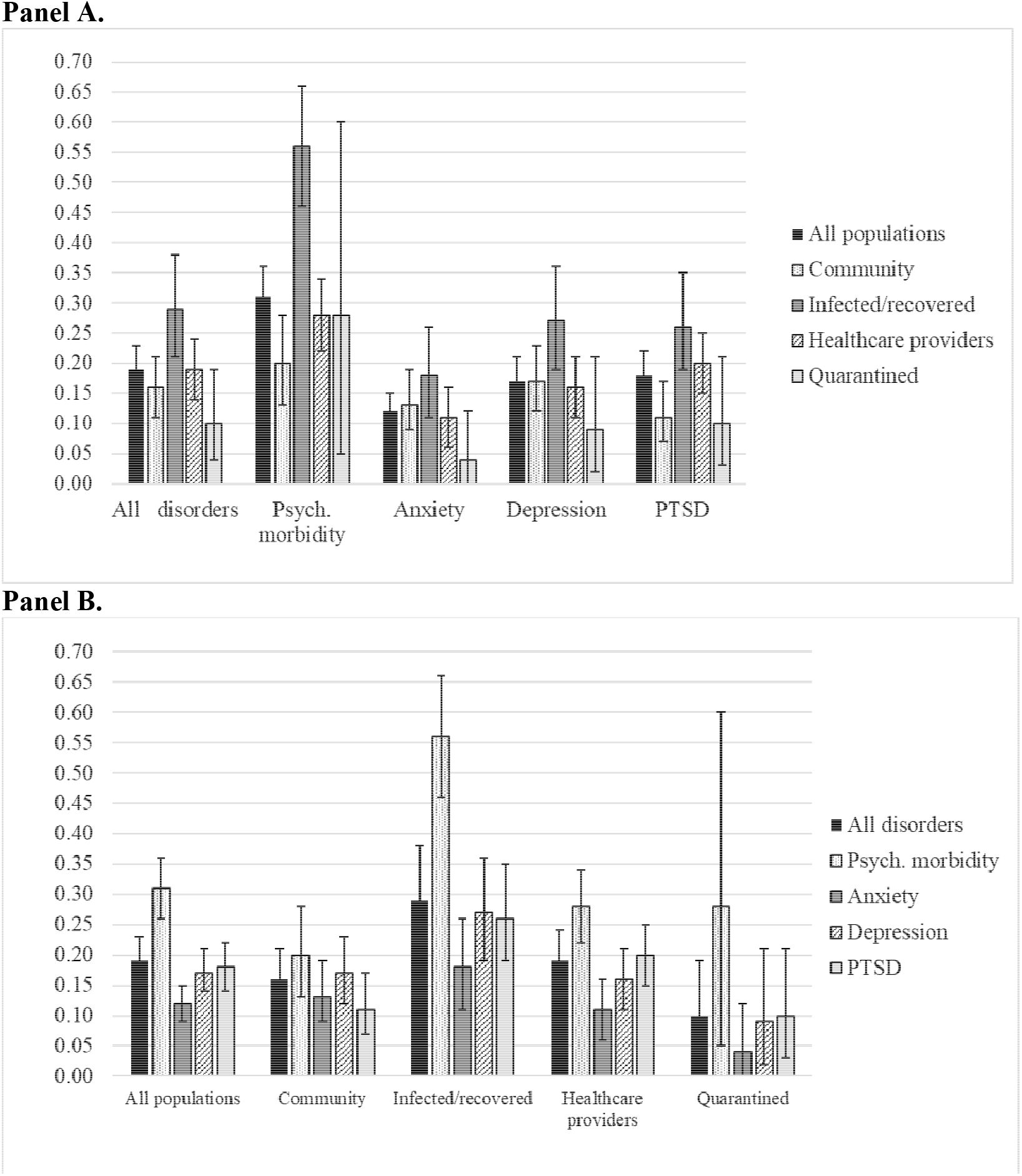
Summary point prevalence estimates and 95% confidence intervals grouped by disorder (Panel A) and population (Panel B).

Across disorders, the highest prevalence was found for infected/recovered adults (29%, 95%CI=21-38%), followed by healthcare providers (19%, 95%CI=14-24%), community adults (16%, 95%CI=11-21%), and quarantined adults (10%, 95%CI=4-19%). The highest prevalence of each disorder was found for infected/recovered adults (18-56%). Healthcare providers had high psychiatric morbidity prevalence (28%, 95%CI=22-34%) and PTSD (20%, 95%CI=15-25%) relative to other disorders. Adults in the community had high prevalence of psychiatric morbidity (20%, 95%CI=13-28%) and depression (17%, 95%CI=12-23%) relative to other disorders. Quarantined adults had much higher prevalence of psychiatric morbidity (28%, 95%CI=5-60%) than other disorders, but summary estimates based on few studies/individual estimates were imprecisely estimated.

In sensitivity analyses, use of the robust estimator resulted in wider confidence intervals on average for summary estimates (Mean Difference=.02, Range= −.64 to .19) and back-transformed point prevalence estimates (Mean Difference=.02, Range= −.55 to .16). Most studies (a) focused on SARS, (b) were conducted in China and (separately) Hong Kong, included estimates (c) measured during the acute phase, and obtained (d) via questionnaire and (e) with standard scoring, and (f) were of low or very low quality (see Table 2). Overall prevalence estimates across disorders and populations were higher for studies/estimates: (1) focused on SARS or MERS versus COVID-19 (22/23% vs. 12%), (2) conducted in the United States, Hong Kong, Korea or Taiwan (23-40%) versus other locations (4-18%), (3) measured post-acute versus acute phase (20/21% versus 18%), obtained (4) via questionnaire versus clinician assessment (20% vs. 10%) and (5) with standard versus non-standard scoring (20% vs. 17%), and (6) of moderate or very low versus low quality (22/20% vs. 17%).

**Table 2.** Sample size and Sensitivity Meta-Analysis/Regression Results, Including Summary Estimates/Prevalence and 95% Confidence Intervals by Pandemic, Location, Timing, Measure, Scoring and Study Quality.

| Factor | Categories | Study *N* | IE *N* | Sample *N* | SE | 95% CI LB | 95% CI UB | SPP | 95% CI LB | 95% CI UB |
| --- | --- | --- | --- | --- | --- | --- | --- | --- | --- | --- |
| Pandemic | COVID-19 | 20 | 287 | 62180 | .37 | .29 | .45 | .12 | .07 | .18 |
|  | MERS | 8 | 24 | 2695 | .49 | .36 | .62 | .22 | .11 | .33 |
|  | SARS | 37 | 497 | 22711 | .50 | .45 | .56 | .23 | .18 | .28 |
| Location | Canada | 5 | 31 | 3858 | .44 | .28 | .61 | .18 | .06 | .32 |
|  | China | 21 | 421 | 52473 | .38 | .31 | .46 | .13 | .08 | .19 |
|  | Hong Kong | 14 | 138 | 6069 | .55 | .45 | .64 | .26 | .18 | .36 |
|  | Korea | 7 | 23 | 2521 | .52 | .38 | .66 | .24 | .13 | .38 |
|  | Saudi Arabia | 1 | 1 | 174 | .22 | -.19 | .63 | .04 | .00 | .34 |
|  | Singapore | 7 | 102 | 4229 | .45 | .32 | .59 | .18 | .09 | .30 |
|  | Taiwan | 7 | 74 | 5977 | .50 | .37 | .64 | .23 | .12 | .35 |
|  | Italy | 1 | 12 | 6873 | .53 | .18 | .88 | .25 | .02 | .59 |
|  | United States | 1 | 3 | 2694 | .69 | .32 | 1.05 | .40 | .09 | .76 |
|  | Multi: Singapore & India | 1 | 3 | 2718 | .24 | -.12 | .61 | .05 | .00 | .32 |
| Timing | Acute | 41 | 436 | 73237 | .45 | .39 | .50 | .18 | .14 | .22 |
|  | Within 1-year post | 16 | 171 | 6286 | .49 | .41 | .57 | .21 | .15 | .28 |
|  | After 1-year post | 12 | 201 | 8063 | .47 | .39 | .56 | .20 | .13 | .27 |
| Measure | Questionnaire | 62 | 696 | 86064 | .47 | .43 | .52 | .20 | .16 | .24 |
|  | Clinician assessment | 9 | 112 | 1522 | .35 | .28 | .41 | .10 | .07 | .15 |
| Scoring | Standard | 55 | 668 | 63186 | .47 | .42 | .51 | .20 | .16 | .24 |
|  | Non-standard | 22 | 140 | 24400 | .43 | .38 | .48 | .17 | .13 | .21 |
| Quality* | Very low | 28 | 297 | 33920 | .47 | .10 | .54 | .20 | .14 | .26 |
|  | Low | 34 | 427 | 49113 | .44 | .38 | .50 | .17 | .13 | .23 |
|  | Moderate | 5 | 84 | 4553 | .50 | .34 | .66 | .22 | .10 | .38 |

\*Two studies with same or overlapping samples but different quality ratings were treated as separate studies. IE = Individual estimate. SE = Summary estimate. CI = Confidence interval. LB = Lower bound. UB = Upper bound. SPP = Summary point prevalence (i.e., back-transformed Freeman-Tukey double arcsine transformed estimates; Miller, 1975).

## 4. Discussion

Across 68 individual studies, about one-in-five adults in populations impacted by a coronavirus pandemic demonstrated or reported signs or symptoms indicative of a mental disorder diagnosis. Undifferentiated psychiatric morbidity was the most prevalent disorder in most of our study populations followed by PTSD and depression, then anxiety. Infected recovered adults were most at risk of a mental disorder, followed by healthcare providers and adults in the community, then quarantined adults. As we discuss below in relation to these populations, our summary estimates tended to be higher, often substantially than 12-month or shorter prevalence estimates from a meta-analysis and systematic review (Somers et al., 2006; Steel et al., 2014) and from epidemiological studies of the adult populations of China (Huang et al., 2019), Europe (ESEMeD/MHEDEA 2000 Investigators, 2004), and the United States (Kessler et al., 2005; Regier et al., 1988). Our results suggest that coronavirus pandemics are associated with an adverse mental health impact as manifested by elevated rates of multiple mental disorders in multiple populations. Our search strategy and analytical methods allowed us to utilize all relevant data from individual studies, while accounting for dependencies in those data. Furthermore, our focus on peer-reviewed studies utilizing standardized measures strengthens our confidence in the validity of our results. Despite these strengths, our results do not provide an indication of causal relations between coronavirus pandemics and mental disorders, as the majority of the observational studies included in our meta-analysis did not include adequate control/comparison groups, and we did not examine related differences.

Infected or recovered adults had the highest prevalence of all disorders and each individual disorder, with notably higher prevalence relative to unselected adults in prior studies (ESEMeD/MHEDEA 2000 Investigators, 2004; Huang et al., 2019; Kessler et al., 2005; Regier et al., 1988). For example, 56% of infected/recovered adults were positive for undifferentiated psychiatric morbidity compared to the 9% (Huang et al., 2019) to 26% of adults (Kessler et al., 2005) who were positive for ‘any disorder’ studies of unselected adults. Prevalence of PTSD (26%) was much higher than found among unselected adults in China (0.2% Huang et al., 2019), Europe (0.9% ESEMeD/MHEDEA 2000 Investigators, 2004), and the United States (3.5%; Kessler et al., 2005), and comparable to *lifetime* prevalence of PTSD among combat veterans (Weiss et al., 1992) and prevalence of PTSD among survivors of intensive care unit treatment (Griffiths et al., 2007). The latter comparison is informative, as the high prevalence of PTSD among infected/recovered adults may be attributable in-part to their experiences of intensive care and associated treatment, such as mechanic ventilation (Bienvenu et al., 2018). Prevalence of depression (27%) was also much higher than the 3-10% prevalence of mood/depressive disorders or major depressive disorder found in studies of unselected adult populations (ESEMeD/MHEDEA 2000 Investigators, 2004; Huang et al., 2019; Kessler et al., 2005; Regier et al., 1988; Steel et al., 2013). Prevalence of anxiety was higher than prevalence found among unselected adults in several studies (e.g., Steel et al., 2013; also see Somers et al., 2006), but identical to unselected adults in the United States (18%; Kessler et al., 2005). Our results suggest that severe infection is a potent risk factor for mental disorders, and as demonstrated by prior studies (e.g., Mak et al., 2009), may contribute to functional impairment and disability that continues to adversely impact the mental health of recovered patients. We hypothesize that even minor infections may be quite threatening and stressful in people with pre-existing physical or mental health conditions. Nuanced investigations of causal links between infection severity, symptom presentation and medical treatments, and mental disorders and facets of (dys)function post recovery will be useful for mental health intervention planning in relation to COVID-19 and future pandemics.

Healthcare providers had high prevalence of most disorders relative to unselected adult population samples (ESEMeD/MHEDEA 2000 Investigators, 2004; Huang et al., 2019; Kessler et al., 2005; Regier et al., 1988; Somers et al., 2006; Steel et al., 2013). Prevalence of undifferentiated psychiatric morbidity (28%) was somewhat higher than found in a study of emergency room physicians and trainees (26.8%; Goh et al., 1999). Prevalence of PTSD (20%) was higher than found in multiple studies of trauma-exposed physicians (see Sendler et al., 2016) and lower than found in a study of critical care nurses (24-29%; Mealer et al., 2007). Prevalence of depression (16%) was higher than found in some (Wurm et al., 2016; Ford et al., 1998), but not other studies of healthcare providers (Frank et al., 1999), though Ford and colleagues (1998) and Frank and colleagues (1999) provided lifetime estimates, which tend to be higher than point- or 12-month prevalence. Notably high PTSD prevalence may reflect exposure to numerous potentially traumatic events (e.g., suffering and death of patients, morally ambiguous decisions and moral injury), whereas job stresses (e.g., long and inflexible hours, fluid work environments and job duties) may contribute to high prevalence of psychiatric morbidity and depression, along with burnout (Goh et al., 1999). Considered together, the results above suggest that healthcare providers are at risk for mental disorders, and especially, traumatic-stress disorders during a coronavirus pandemic. Future observational research testing this hypothesis will benefit from case-control and cohort designs that more clearly establish causal links between particular stressors, threats and traumas experienced by healthcare providers, and development of various mental disorders.

Prevalence of depression (17%) and PTSD (11%) was notably elevated among adults in the community relative to those found among samples of unselected adult (ESEMeD/MHEDEA 2000 Investigators, 2004; Huang et al., 2019; Kessler et al., 2005; Regier et al., 1988; Somers et al., 2006; Steel et al., 2013). Prevalence of anxiety (13%) and undifferentiated psychiatric morbidity (20%) was higher than comparable rates found in some (ESEMeD/MHEDEA 2000 Investigators, 2004; Huang et al., 2019; Steel et al., 2013), but not other studies of unselected adult samples (Kessler et al., 2005). These results suggest coronavirus pandemics have a nuanced impact on adults in the community. The high prevalence of depression, in particular, might reflect the stress of living and coping with the threat of infection, societal dysfunction, and/or limited availability of routines and resources. Thus, countries that continue to be impacted by COVID-19 may experience increased incidence of depression in the general population over time. Longitudinal studies including adequate control/comparison groups will be useful in testing this and related hypotheses.

Quarantined adults had rates of undifferentiated psychiatric morbidity (28%), anxiety (4%) and depression (9%) that were higher than some studies of unselected adults (Huang et al., 2019; Regier et al., 1988), and lower than others (Kessler et al., 2005). Prevalence of PTSD (10%), on the other hand, was higher than found in multiple studies of unselected adults (ESEMeD/MHEDEA 2000 Investigators, 2004; Huang et al., 2019; Kessler et al., 2005). Yet, these findings must be interpreted more cautiously than findings for other populations given the small number of studies/estimates from which summary estimates were derived. As a front-line defense against coronavirus pandemics with substantial benefits as well as costs, quarantine draws advocates and critics in equal measure. Additional research is needed, especially to investigate for whom and under what conditions quarantine is associated with mental disorders. There a likely multiple mediators and moderators of links between quarantine and mental disorders, including degree of restriction/isolation (e.g., shelter-in-place, isolation on an intensive care unit), demographic factors (age, sex, relationship status, parental status), and supports that mitigate the impact of quarantine (e.g., food delivery to people with limited mobility). Future research that provides rigorous and actionable data can facilitate effective responding by communities and countries.

Several issues should be considered when interpreting the results of this meta-analysis. First, as indicated by large CI, some studies produced imprecise summary prevalence estimates, likely due to small sample sizes. Second, most studies were of low or very low quality. Summary estimates varied somewhat by study quality, though summary estimates were identical for moderate and very low quality studies. Third, the true prevalence of diagnosable mental disorders in populations impacted by coronavirus epidemics is likely to be lower than the magnitude of summary estimates, derived mostly from studies using self-report questionnaires that do not precisely assess mental disorder diagnostic criteria. Only a subset of included studies utilized gold-standard clinical interviews (e.g., Structured Clinical Interview for DSM) and assessment instruments specifically designed to screen for mental disorders (e.g., Kessler et al., 2002). Indeed, sensitivity analyses revealed higher overall summary prevalence obtained from questionnaires versus clinician assessment. Somewhat mitigating this concern: (a) almost all studies utilizing questionnaires implemented empirically validated cutoffs to identify participants with mental disorders (e.g., Plummer et al., 2016); and (b) studies that did not implement standard questionnaire cutoffs (and those that utilized unknown scoring algorithms to identify mental disorders) yielded similar overall summary prevalence estimates as those that did.

As applied to understanding and planning for COVID-19 (Boden et al., 2020), summary estimates must be interpreted cautiously. Overall prevalence of all disorders across all populations was lower for COVID-19 versus MERS and SARS. SARS and MERS have higher mortality rates and higher average severity than does COVID-19. The greater threat to health and mortality of SARS and MERS, and the challenges of long-term recovery from severe infection may have potentially increased risk for mental disorders among those infected with and recovered from these viruses, healthcare workers treating infected patients, and the general public who worried about severe infection, disability and death. Also potentially accounting for differences, all COVID-19 studies focused on mental disorder prevalence during the acute phase of the pandemic, whereas MERS and SARS studies included acute and longer time-frames, thus capturing mental dysfunction across a longer period of time. Yet, the overall mental disorder prevalence varied little between acute and longer time-frames. In relation to COVID-19, it will be important to investigate whether mental disorder prevalence increases in countries with prolonged periods of infection and economic hardship (e.g., United States, Brazil). Further suggesting the need for such investigations, our sensitivity analyses demonstrated differences in prevalence by country, with the United States, Hong Kong, Korea and Taiwan having notably higher prevalence than Canada, Singapore, Taiwan, China and Saudi Arabia. Caution is advised when comparing prevalence rates by country, as estimates were derived from a varying number of studies for each country, and a single study provided estimates for some countries (e.g., United States). Regardless, we hypothesize that countries and areas within countries that experience prolonged infection, quarantine and economic hardship will experience increased rates of the disorders examined in this study, in addition to disorders of despair (e.g., alcohol dependence; Petterson et al., 2020) and related to disease threat (e.g., illness anxiety disorder, somatic symptom disorder). The raw number of mental disorders attributable to COVID-19 will be substantial and magnitudes higher than attributable to MERS and SARS due to the vast scope and ongoing nature of the pandemic.

Preliminary evidence suggests that COVID-19 exacerbates existing inequalities that harm racial/ethnic minorities, economically disadvantaged people, people with ‘essential’ job types, etc. (Hooper et al., 2020; van Dorn et al., 2020). Additional research is needed to quantify the mental health impact of COVID-19 in these subpopulations, and to document disparities in assessment and treatment of COVID-19-related mental disorders. Future research that examines the potentially manifold pathways to individual outcomes among subpopulations most at-risk will be instrumental in intervening in a cost-effective, effective and equitable manner. Future research that utilizes rigorous and validated subject recruitment and mental disorder assessment methods can improve upon the low quality of the majority of studies included in our study. Additional limitations can be addressed through assessment of pre-existing mental disorders among study samples, rigorous assessment of time-periods at risk, and/or inclusion of control conditions (e.g., people exposed to but not infected by COVID-19 as a comparator for infected/recovered people). Studies that do so will more directly measure the impact of COVID-19-related threats, stressors and traumas on mental disorder prevalence, incidence, and rate ratios, which will be essential to intervention planning and implementation.

Comparison of our synthesized prevalence rates across populations and disorder demonstrate the nuanced mental health impact of coronavirus pandemics. These data may prove a useful guide for future research and data collection that results in actionable information that can be utilized in regard to mental health treatment and resource allocation and policy. In addition to examining specific disorders in targeted and vulnerable populations, this research should examine between-country, and even, between-community and neighborhood variation that results from the confluence of specific threats, stressors and traumas prevalent in a given area.

## Data Availability

Data is available upon request of the first author.

## References

* Study included in meta-analysis.

* Al-Rabiaah A, Temsah MH, Al-Eyadhy AA, Hasan GM, Al-Zamil F, Al-Subaie S, Alsohime F, Jamal A, Alhaboob A, Al-Saadi B, Somily AM. Middle East Respiratory Syndrome-Corona Virus (MERS-CoV) associated stress among medical students at a university teaching hospital in Saudi Arabia. Journal of Infection & Public Health. 2020; 13(5):687–691 doi:10.1016/j.jiph.2020.01.005

3. Anderson RJ, Freedland KE, Clouse RE, Lustman PJ. The prevalence of comorbid depression in adults with diabetes: a meta-analysis. Diabetes care. 2001;24(6):1069–78. doi: 10.2337/diacare.24.6.1069.

4. Bai Y, Lin CC, Lin CY, Chen JY, Chue CM, Chou P. Survey of stress reactions among health care workers involved with the SARS outbreak. Psychiatric Services. 2004;55(9):1055–7. doi:10.1176/appi.ps.55.9.1055

5. Bienvenu OJ, Friedman LA, Colantuoni E, Dinglas VD, Sepulveda KA, Mendez-Tellez P, Shanholz C, Pronovost PJ, Needham DM. Psychiatric symptoms after acute respiratory distress syndrome: a 5-year longitudinal study. Intensive Care Medicine. 2018;44(1):38–47. doi:10.1007/s00134-017-5009-4

6. Boden M. Estimating the mental health impact of COVID19 on United States Populations. 2020. [cited 2020 Sep 19]; Available from: https://osf.io/ytd92

7. Brooks SK, Webster RK, Smith LE, Woodland L, Wessely S, Greenberg N, Rubin GJ. The psychological impact of quarantine and how to reduce it: rapid review of the evidence. The Lancet. 2020; 395:912–20. doi:10.1016/S0140-6736(20)30460-8

* Cao W, Fang Z, Hou G, Han M, Xu X, Dong J, Zheng J. The psychological impact of the COVID-19 epidemic on college students in China. Journal of Psychiatric Research. 2020:287(112934)1–5. doi:10.1016/j.psychres.2020.112934

* Casagrande M, Favieri F, Tambelli R, Forte G. The enemy who sealed the world: Effects quarantine due to the COVID-19 on sleep quality, anxiety, and psychological distress in the Italian population. Sleep Medicine. 2020;75:12–20. doi: 10.1016/j.sleep.2020.05.011

* Chan AO, Huak CY. Psychological impact of the 2003 severe acute respiratory syndrome outbreak on health care workers in a medium size regional general hospital in Singapore. Occupational Medicine. 2004;54(3):190–6. doi:10.1093/occmed/kqh027

* Chan SS, So WK, Wong DC, Lee AC, Tiwari A. Improving older adults’ knowledge and practice of preventive measures through a telephone health education during the SARS epidemic in Hong Kong: A pilot study. International Journal of Nursing Studies. 2007;44(7):1120–7. doi:10.1016/j.ijnurstu.2006.04.019

* Chen CS, Wu HY, Yang P, Yen CF. Psychological distress of nurses in Taiwan who worked during the outbreak of SARS. Psychiatric Services. 2005;56(1):76–9. doi:10.1176/appi.ps.56.1.76

* Cheng SK, Wong CW, Tsang J, Wong KC. Psychological distress and negative appraisals in survivors of severe acute respiratory syndrome (SARS). Psychological Medicine. 2004;34(7):1187. doi:10.1017/S0033291704002272

* Chew NW, Lee GK, Tan BY, Jing M, Goh Y, Ngiam NJ, Yeo LL, Ahmad A, Khan FA, Shanmugam GN, Sharma AK. A multinational, multicentre study on the psychological outcomes and associated physical symptoms amongst healthcare workers during COVID-19 outbreak. Brain, Behavior, and Immunity. 2020;88:559–65. doi: 10.1016/j.bbi.2020.04.049

* Chong MY, Wang WC, Hsieh WC, Lee CY, Chiu NM, Yeh WC, Huang TL, Wen JK, Chen CL. Psychological impact of severe acute respiratory syndrome on health workers in a tertiary hospital. British Journal of Psychiatry. 2004;185(2):127–33. doi:10.1192/bjp.185.2.127

16. Cisler JM, Koster EH. Mechanisms of attentional biases towards threat in anxiety disorders: An integrative review. Clinical Psychology Review. 2010;30(2):203–16. doi:10.1016/j.cpr.2009.11.003

17. de Pablo GS, Serrano JV, Catalan A, Arango C, Moreno C, Ferre F, Shin JI, Sullivan S, Brondino N, Solmi M, Fusar-Poli P. Impact of coronavirus syndromes on physical and mental health of health care workers: Systematic review and meta-analysis. Journal of Affective Disorders. 2020;275:48–57. doi:10.1016/j.jad.2020.06.022

18. Dooley D, Catalano R, Wilson G. Depression and unemployment: panel findings from the Epidemiologic Catchment Area study. American Journal of Community Psychology. 1994;22(6):745–65. doi:10.1007/BF02521557

19. ESEMeD/MHEDEA 2000 Investigators, Alonso J, Angermeyer MC, Bernert S, Bruffaerts R, Brugha TS, Bryson H, de Girolamo G, de Graaf R, Demyttenaere K, Gasquet I. Prevalence of mental disorders in Europe: results from the European Study of the Epidemiology of Mental Disorders (ESEMeD) project. Acta Psychiatrica Scandinavica. 2004;109:21–7. doi:10.1111/j.1600-0047.2004.00327

20. Ford DE, Mead LA, Chang PP, Cooper-Patrick L, Wang NY, Klag MJ. Depression is a risk factor for coronary artery disease in men: the precursors study. Archives of Internal Medicine. 1998;158(13):1422–6. doi: 10.1001/archinte.158.13.1422

21. Frank E, Dingle AD. Self-reported depression and suicide attempts among US women physicians. American Journal of Psychiatry. 1999;156(12):1887–94. doi: 10.1176/ajp.156.12.1887

22. Freeman MF, Tukey JW. Transformations related to the angular and the square root. The Annals of Mathematical Statistics. 1950;21(4):607–11. doi:10.1214/aoms/1177729756

23. Galea S, Merchant RM, Lurie N. The mental health consequences of COVID-19 and physical distancing: The need for prevention and early intervention. JAMA Internal Medicine. 2020;180(6):817–8. doi:10.1001/jamainternmed.2020.1562

24. Garfin DR, Silver RC, Holman EA. The novel coronavirus (COVID-2019) outbreak: Amplification of public health consequences by media exposure. Health Psychology. 2020;39(5):355–7. doi: 10.1037/hea0000875

25. Goh L, Cameron PA, Mark P. Burnout in emergency physicians and trainees in Australasia. Emergency Medicine. 1999;11(4):250–7. doi: 10.1046/j.1442-2026.1999.00071.x

26. GRADE Working Group. Grading quality of evidence and strength of recommendations. BMC. 2004;328(7454):1490. doi:10.1136/bmj.328.7454.1490

27. Griffiths J, Fortune G, Barber V, Young JD. The prevalence of post traumatic stress disorder in survivors of ICU treatment: a systematic review. Intensive Care Medicine. 2007;33(9):1506–18. doi:10.1007/s00134-007-0730-z

* Guo Q, Zheng Y, Shi J, Wang J, Li G, Li C, Fromson JA, Xu Y, Liu X, Xu H, Zhang T. Immediate psychological distress in quarantined patients with COVID-19 and its association with peripheral inflammation: a mixed-method study. Brain, Behavior, and Immunity. 20201;88:17–27. doi: 10.1016/j.bbi.2020.05.038

29. Hammen C. Stress and depression. Annual Review of Clinical Psychology. 2005;1:293–319. doi:10.1146/annuerev.clinpsy.1.102803.143938

* Hao F, Tan W, Jiang L, Zhang L, Zhao X, Zou Y, Hu Y, Luo X, Jiang X, McIntyre RS, Tran B. Do psychiatric patients experience more psychiatric symptoms during COVID-19 pandemic and lockdown? A case-control study with service and research implications for immunopsychiatry. Brain, Behavior & Immunity. 2020;87:100–6. doi:10.1016/j.bbi.2020.04.069

* Hawryluck L, Gold WL, Robinson S, Pogorski S, Galea S, Styra R. SARS control and psychological effects of quarantine, Toronto, Canada. Emerging Infectious Diseases. 2004;10(7):1206. doi:10.3201/eid1007.030703

32. Hedges LV, Tipton E, Johnson MC. Robust variance estimation in metalJregression with dependent effect size estimates. Research Synthesis Methods. 2010;1(1):39–65. doi:10.1002/jrsm.5

* Hong X, Currier GW, Zhao X, Jiang Y, Zhou W, Wei J. Posttraumatic stress disorder in convalescent severe acute respiratory syndrome patients: a 4-year follow-up study. General Hospital Psychiatry. 2009;31(6):546–54. doi:10.1016/j.genhosppsych.2009.06.008

34. Hooper MW, Nápoles AM, Pérez-Stable EJ. COVID-19 and racial/ethnic disparities. JAMA. 2020;323(24):2466–7. doi:10.1001/jama.2020.8598

35. Huang Y, Wang Y, Wang H, Liu Z, Yu X, Yan J, Yu Y, Kou C, Xu X, Lu J, Wang Z. Prevalence of mental disorders in China: a cross-sectional epidemiological study. The Lancet Psychiatry. 2019;6(3):211–24. doi:10.1016/S2215-0366(18)30511-X

* Huang Y, Zhao N. Generalized anxiety disorder, depressive symptoms and sleep quality during COVID-19 outbreak in China: a web-based cross-sectional survey. Psychiatry Research. 2020;288(112954):1–6. doi:10.1016/j.psychres.2020.112954

* Jeong H, Yim HW, Song YJ, Ki M, Min JA, Cho J, Chae JH. Mental health status of people isolated due to Middle East Respiratory Syndrome. Epidemiological Health. 2016;38. doi:10.4178/epih.e2016048

* Jung H, Jung SY, Lee MH, Kim MS. Assessing the Presence of Post-Traumatic Stress and Turnover Intention Among Nurses Post–Middle East Respiratory Syndrome Outbreak: The Importance of Supervisor Support. Workplace Health Safety. 2020;68(7):337–45. doi:10.1177/2165079919897693

39. Kessler RC, Andrews G, Colpe LJ, Hiripi E, Mroczek DK, Normand SL, Walters EE, Zaslavsky AM. Short screening scales to monitor population prevalences and trends in non-specific psychological distress. Psychological medicine. 2002;32(6):959–76. doi:10.1017/S0033291702006074

40. Kessler RC, Chiu WT, Demler O, Walters EE. Prevalence, severity, and comorbidity of 12-month DSM-IV disorders in the National Comorbidity Survey Replication. Archives of general psychiatry. 2005;62(6):617–27. doi: 10.1001/archpsyc.62.6.617

* Kim HC, Yoo SY, Lee BH, Lee SH, Shin HS. Psychiatric findings in suspected and confirmed Middle East respiratory syndrome patients quarantined in hospital: a retrospective chart analysis. Psychiatry Investigation. 2018;15(4):355. doi:10.30773/pi.2017.10.25.1

* Kim Y, Seo E, Seo Y, Dee V, Hong E. Effects of Middle East Respiratory Syndrome Coronavirus on post-traumatic stress disorder and burnout among registered nurses in South Korea. International Journal of Healthcare. 2018;4(2):27–33. doi:10.5430/ijh.v4n2p27

* Kwek SK, Chew WM, Ong KC, Ng AW, Lee LS, Kaw G, Leow MK. Quality of life and psychological status in survivors of severe acute respiratory syndrome at 3 months postdischarge. Journal of Psychosomatic Research. 2006;60(5):513–9. doi:10.1016/j.jpsychores.2005.08.020

* Lai J, Ma S, Wang Y, Cai Z, Hu J, Wei N, Wu J, Du H, Chen T, Li R, Tan H. Factors associated with mental health outcomes among health care workers exposed to coronavirus disease 2019. JAMA Network Open. 2020;3(3):e203976.. doi:10.1001/jamanetworkopen.2020.3976

* Lam MH, Wing YK, Yu MW, Leung CM, Ma RC, Kong AP, So WY, Fong SY, Lam SP. Mental morbidities and chronic fatigue in severe acute respiratory syndrome survivors: long-term follow-up. Archives of Internal Medicine. 2009;169(22):2142–7. doi:10.1001/archinternmed.2009.384

* Lancee WJ, Maunder RG, Goldbloom DS. Prevalence of psychiatric disorders among Toronto hospital workers one to two years after the SARS outbreak. Psychiatry Services. 2008;59(1):91–5. doi:10.1176/ps.2008.59.1.91

* Lau JT, Yang X, Tsui HY, Pang E, Wing YK. Positive mental health-related impacts of the SARS epidemic on the general public in Hong Kong and their associations with other negative impacts. Journal of Infection. 2006;53(2):114–24. doi:10.1016/j.jinf.2005.10.019

* Lee AM, Wong JG, McAlonan GM, Cheung V, Cheung C, Sham PC, Chu CM, Wong PC, Tsang KW, Chua SE. Stress and psychological distress among SARS survivors 1 year after the outbreak. The Canadian Journal of Psychiatry. 2007;52(4):233–40. doi:10.1177/070674370705200405

* Lee DT, Sahota D, Leung TN, Yip AS, Lee FF, Chung TK. Psychological responses of pregnant women to an infectious outbreak: a case-control study of the 2003 SARS outbreak in Hong Kong. Journal of Psychosomatic Research. 2006;61(5):707–13. doi:10.1016/j.jpsychores.2006.08.005

* Lee SH, Shin HS, Park HY, Kim JL, Lee JJ, Lee H, Won SD, Han W. Depression as a mediator of chronic fatigue and post-traumatic stress symptoms in Middle East respiratory syndrome survivors. Psychiatry Investigation. 2019;16(1):59. doi:10.30773/pi.2018.10.22.3

* Lee SM, Kang WS, Cho AR, Kim T, Park JK. Psychological impact of the 2015 MERS outbreak on hospital workers and quarantined hemodialysis patients. Comprehensive Psychiatry. 2018;87:123–7. doi:10.1016/j.comppsych.2018.10.003

* Lee TM, Chi I, Chung LM, Chou KL. Ageing and psychological response during the post-SARS period. Aging and Mental Health. 2006;10(3):303–11. doi:10.1080/13607860600638545

* Leung GM, Lam TH, Ho LM, Ho SY, Chan BH, Wong IO, Hedley AJ. The impact of community psychological responses on outbreak control for severe acute respiratory syndrome in Hong Kong. Journal of Epidemiology & Community Health. 2003;57(11):857–63. doi:10.1136/jech.57.11.857

* Lin CY, Peng YC, Wu YH, Chang J, Chan CH, Yang DY. The psychological effect of severe acute respiratory syndrome on emergency department staff. Emergency Medicine Journal. 2007;24(1):12–7. doi:10.1136/emj.2006.035089

* Liu CH, Zhang E, Wong GT, Hyun S. Factors associated with depression, anxiety, and PTSD symptomatology during the COVID-19 pandemic: Clinical implications for US young adult mental health. Psychiatry Research. 20201;290:113172. doi: 10.1016/j.psychres.2020.113172

* Liu N, Zhang F, Wei C, Jia Y, Shang Z, Sun L, Wu L, Sun Z, Zhou Y, Wang Y, Liu W. Prevalence and predictors of PTSS during COVID-19 outbreak in China hardest-hit areas: Gender differences matter. Psychiatry Research. 2020;287(112921):1–7. doi:10.1016/j.psychres.2020.112921

* Liu X, Kakade M, Fuller CJ, Fan B, Fang Y, Kong J, Guan Z, Wu P. Depression after exposure to stressful events: lessons learned from the severe acute respiratory syndrome epidemic. Comprehensive Psychiatry. 2012;53(1):15–23. doi:10.1016/j.comppsych.2011.02.003

* Lu W, Wang H, Lin Y, Li L. Psychological status of medical workforce during the COVID-19 pandemic: A cross-sectional study. Psychiatry Research. 2020;288(112936):1–5. doi:10.1016/j.psychres.2020.112936

* Lung F, Lu YC, Chang YY, Shu BC. Mental symptoms in different health professionals during the SARS attack: A follow-up study. Psychiatric Quarterly. 2009;80(2):107. doi:10.1007/s11126-009-9095-5

* Mak IW, Chu CM, Pan PC, Yiu MG, Chan VL. Long-term psychiatric morbidities among SARS survivors. General Hospital Psychiatry. 2009;31(4):318–26. doi:10.1016/j.genhosppsych.2009.03.001

* Mak IW, Chu CM, Pan PC, Yiu MG, Ho SC, Chan VL. Risk factors for chronic post-traumatic stress disorder (PTSD) in SARS survivors. General Hospital Psychiatry. 2010;32(6):590–8. doi:10.1016/j.genhosppsych.2010.07.007

* Maunder RG, Lancee WJ, Balderson KE, Bennett JP, Borgundvaag B, Evans S, Fernandes CM, Goldbloom DS, Gupta M, Hunter JJ, Hall LM. Long-term psychological and occupational effects of providing hospital healthcare during SARS outbreak. Emerging Infectious Diseases. 2006;12(12):1924. doi:10.3201/eid1212.060584

63. Mealer ML, Shelton A, Berg B, Rothbaum B, Moss M. Increased prevalence of post-traumatic stress disorder symptoms in critical care nurses. American journal of respiratory and critical care medicine. 2007;175(7):693–7. doi: 10.1164/rccm.200606-735OC

* Mihashi M, Otsubo Y, Yinjuan X, Nagatomi K, Hoshiko M, Ishitake T. Predictive factors of psychological disorder development during recovery following SARS outbreak. Health Psychology. 2009;28(1):91. doi:10.1037/a0013674

65. Miller JJ. The inverse of the Freeman–Tukey double arcsine transformation. The American Statistician. 1978;32(4):138. doi:10.1080/00031305.1978.10479283

* Moccia L, Janiri D, Pepe M, Dattoli L, Molinaro M, De Martin V, Chieffo D, Janiri L, Fiorillo A, Sani G, Di Nicola M. Affective temperament, attachment style, and the psychological impact of the COVID-19 outbreak: an early report on the Italian general population. Brain, Behavior & Immunity. 2020;87:75–79. doi:10.1016/j.bbi.2020.04.048

67. Moher D, Liberati A, Tetzlaff J, Altman DG, Prisma Group. Preferred reporting items for systematic reviews and meta-analyses: the PRISMA statement. PLoS Medicine. 2009;6(7):e1000097. doi:10.1371/journal.pmed.1000097

* Moldofsky H, Patcai J. Chronic widespread musculoskeletal pain, fatigue, depression and disordered sleep in chronic post-SARS syndrome; a case-controlled study. BMC Neurology. 2011;11(1):37. doi:10.1186/1471-2377-11-37

* Nickell LA, Crighton EJ, Tracy CS, Al-Enazy H, Bolaji Y, Hanjrah S, Hussain A, Makhlouf S, Upshur RE. Psychosocial effects of SARS on hospital staff: survey of a large tertiary care institution. Canadian Medical Association Journal. 2004;170(5):793–8. doi:10.1503/cmaj.1031077

* Peng EY, Lee MB, Tsai ST, Yang CC, Morisky DE, Tsai LT, Weng YL, Lyu SY. Population-based post-crisis psychological distress: an example from the SARS outbreak in Taiwan. Journal of the Formosan Medical Association. 2010;109(7):524–32. doi:10.1016/S0929-6646(10)60087-3

71. Petterson S, Westfall JM, Miller BF. Projected Deaths of Despair from COVID-19. Well Being Trust. 2020. [cited 2020 Sep 19]; Available from: https://wellbeingtrust.org/wp-content/uploads/2020/05/WBT_Deaths-of-Despair_COVID-19-FINAL-FINAL.pdf

72. Plummer F, Manea L, Trepel D, McMillan D. Screening for anxiety disorders with the GAD-7 and GAD-2: A systematic review and diagnostic metaanalysis. General Hospital Psychiatry. 2016 Mar 1;39:24–31. doi:10.1016/j.genhosppsych.2015.11.005

73. Powers MB, Halpern JM, Ferenschak MP, Gillihan SJ, Foa EB. A meta-analytic review of prolonged exposure for posttraumatic stress disorder. Clinical Psychology Review. 2010;30(6):635–41. doi:10.1016/j.cpr.2010.04.007

74. Regier DA, Boyd JH, Burke JD, Rae DS, Myers JK, Kramer M, Robins LN, George LK, Karno M, Locke BZ. One-month prevalence of mental disorders in the United States: Based on five epidemiologic catchment area sites. Archives of General Psychiatry. 1988;45(11):977–86. doi:10.1001/archpsyc.1988.01800350011002

* Reynolds DL, Garay JR, Deamond SL, Moran MK, Gold W, Styra R. Understanding, compliance and psychological impact of the SARS quarantine experience. Epidemiology & Infection. 2008;136(7):997–1007. doi:10.1017/S0950268807009156

76. Rogers JP, Chesney E, Oliver D, Pollak TA, McGuire P, Fusar-Poli P, et al. Psychiatric and neuropsychiatric presentations associated with severe coronavirus infections: a systematic review and meta-analysis with comparison to the COVID-19 pandemic. The Lancet Psychiatry. 2020;7(7):611–27. doi:10.1016/S2215-0366(20)30203-0

77. Ross LE, Grigoriadis S, Mamisashvili L, Koren G, Steiner M, Dennis CL, Cheung A, Mousmanis P. Quality assessment of observational studies in psychiatry: an example from perinatal psychiatric research. International Journal of Methods in Psychiatric Research. 2011;20(4):224–34. doi:10.1002/mpr.356

78. Sendler DJ, Rutkowska A, Makara-Studzinska M. How the exposure to trauma has hindered physicians’ capacity to heal: prevalence of PTSD among healthcare workers. The European Journal of Psychiatry. 2016;30(4):321–34.

* Sheng B, Cheng SK, Lau KK, Li HL, Chan EL. The effects of disease severity, use of corticosteroids and social factors on neuropsychiatric complaints in severe acute respiratory syndrome (SARS) patients at acute and convalescent phases. European Psychiatry. 2005;20(3):236–42. doi:10.1016/j.eurpsy.2004.06.023

* Sim K, Chan YH, Chong PN, Chua HC, Soon SW. Psychosocial and coping responses within the community health care setting towards a national outbreak of an infectious disease. Journal of Psychosomatic Research. 2010;68(2):195–202. doi:10.1016/j.jpsychores.2009.04.004

* Sim K, Phui NC, Yiong HC, Soon WS. Severe acute respiratory syndrome-related psychiatric and posttraumatic morbidities and coping responses in medical staff within a primary health care setting in Singapore. Journal of Clinical Psychiatry. 2004;65(8). doi:10.4088/JCP.v65n0815

* Sin SS, Huak CY. Psychological impact of the SARS outbreak on a Singaporean rehabilitation department. International Journal of Therapy and Rehabilitation. 2004;11(9):417–24. doi:10.12968/ijtr.2004.11.9.19589

83. Somers JM, Goldner EM, Waraich P, Hsu L. Prevalence and incidence studies of anxiety disorders: a systematic review of the literature. The Canadian Journal of Psychiatry. 2006;51(2):100–13. doi: 10.1177/070674370605100206

* Son H, Lee WJ, Kim HS, Lee KS, You M. Hospital workers’ psychological resilience after the 2015 Middle East respiratory syndrome outbreak. Social Behavior and Personality: An International Journal. 2019;47(2):1–3. doi:10.2224/sbp.7228

85. Steel Z, Marnane C, Iranpour C, Chey T, Jackson JW, Patel V, Silove D. The global prevalence of common mental disorders: a systematic review and meta-analysis 1980–2013. International Journal of Epidemiology. 2014;43(2):476–93. doi:10.1093/ije/dyu038

86. Stroup DF, Berlin JA, Morton SC, Olkin I, Williamson GD. MOOSE guidelines for meta-analyses and systematic reviews of observational studies. JAMA. 2000;283(15):2008–12.

* Su TP, Lien TC, Yang CY, Su YL, Wang JH, Tsai SL, Yin JC. Prevalence of psychiatric morbidity and psychological adaptation of the nurses in a structured SARS caring unit during outbreak: a prospective and periodic assessment study in Taiwan. Journal of Psychiatric Research. 2007;41(1-2):119–30. doi:10.1016/j.jpsychires.2005.12.006

* Tam CW, Pang EP, Lam LC, Chiu HF. Severe acute respiratory syndrome (SARS) in Hong Kong in 2003: stress and psychological impact among frontline healthcare workers. Psychological Medicine. 2004;34(7):1197. doi:10.1017/S0033291704002247

* Tan W, Hao F, McIntyre RS, Jiang L, Jiang X, Zhang L, Zhao X, Zou Y, Hu Y, Luo X, Zhang Z. Is returning to work during the COVID-19 pandemic stressful? A study on immediate mental health status and psychoneuroimmunity prevention measures of Chinese workforce. Brain, Behavior, and Immunity. 2020;87:84–92. doi: 10.1016/j.bbi.2020.04.055

* Tang W, Hu T, Hu B, Jin C, Wang G, Xie C, Chen S, Xu J. Prevalence and correlates of PTSD and depressive symptoms one month after the outbreak of the COVID-19 epidemic in a sample of home-quarantined Chinese university students. Journal of Affective Disorders. 2020;274:1–7. doi: 10.1016/j.jad.2020.05.009

* Tham KY, Tan YH, Loh OH, Tan WL, Ong MK, Tang HK. Psychological morbidity among emergency department doctors and nurses after the SARS outbreak. Hong Kong Journal of Emergency Medicine. 2005;12(4):215–23. doi:10.1177/102490790501200404

92. van Dorn A, Cooney RE, Sabin ML. COVID-19 exacerbating inequalities in the US. Lancet (London, England). 2020;395(10232):1243. doi:10.1016/S0140-6736(20)30893-X

* Verma S, Mythily S, Chan YH, Deslypere JP, Teo EK, Chong SA. Post-SARS psychological morbidity and stigma among general practitioners and traditional Chinese medicine practitioners in Singapore. Annals of the Academy of Medicine, Singapore. 2004;33(6):743-8. PMID:15608831

94. Viechtbauer W. Conducting meta-analyses in R with the metafor package. Journal of Statistical Software. 2010;36(3):1–48. doi:10.18637/jss.v036.i03

* Wang C, Pan R, Wan X, Tan Y, Xu L, Ho CS, Ho RC. Immediate psychological responses and associated factors during the initial stage of the 2019 coronavirus disease (COVID-19) epidemic among the general population in China. International Journal of Environmental Research and Public Health. 2020;17(5):1729. doi:10.3390/ijerph17051729

* Wang Y, Di Y, Ye J, Wei W. Study on the public psychological states and its related factors during the outbreak of coronavirus disease 2019 (COVID-19) in some regions of China. Psychology, Health & Medicine. 2020. doi:10.1080/13548506.2020.1746817

97. Weiss DS, Marmar CR, Schlenger WE, Fairbank JA, Kathleen Jordan B, Hough RL, Kulka RA. The prevalence of lifetime and partial postlJtraumatic stress disorder in Vietnam theater veterans. Journal of Traumatic Stress. 1992;5(3):365–76. doi:10.1002/jts.2490050304

* Wu KK, Chan SK, Ma TM. Posttraumatic stress after SARS. Emerging Infectious Diseases. 2005;11(8):1297. doi:10.3201/eid1108.041083

* Wu KK, Chan SK, Ma TM. Posttraumatic stress, anxiety, and depression in survivors of severe acute respiratory syndrome (SARS). Journal of Traumatic Stress: Official Publication of The International Society for Traumatic Stress Studies. 2005;18(1):39–42. doi:10.1002/jts.20004

* Wu P, Fang Y, Guan Z, Fan B, Kong J, Yao Z, Liu X, Fuller CJ, Susser E, Lu J, Hoven CW. The psychological impact of the SARS epidemic on hospital employees in China: exposure, risk perception, and altruistic acceptance of risk. The Canadian Journal of Psychiatry. 2009;54(5):302–11. doi:10.1177/070674370905400504

* Wu P, Liu X, Fang Y, Fan B, Fuller CJ, Guan Z, Yao Z, Kong J, Lu J, Litvak IJ. Alcohol abuse/dependence symptoms among hospital employees exposed to a SARS outbreak. Alcohol & Alcoholism. 2008;43(6):706–12. doi:10.1093/alcalc/agn073

102. Wurm W, Vogel K, Holl A, Ebner C, Bayer D, Mörkl S, Szilagyi IS, Hotter E, Kapfhammer HP, Hofmann P. Depression-burnout overlap in physicians. PloS one. 2016;11(3):e0149913. doi: 10.1371/journal.pone.0149913

* Yin Q, Sun Z, Liu T, Ni X, Deng X, Jia Y, Shang Z, Zhou Y, Liu W. Posttraumatic stress symptoms of health care workers during the corona virus disease 2019. Clinical Psychology & Psychotherapy. 2020;27(3):384–95. doi: 10.1002/cpp.2477

* Yu HY, Ho SC, So KF, Lo YL. The psychological burden experienced by Hong Kong midlife women during the SARS epidemic. Stress and health. 2005;21(3):177–84. doi:10.1002/smi.1051

* Zhang WR, Wang K, Yin L, Zhao WF, Xue Q, Peng M, Min BQ, Tian Q, Leng HX, D. JL, Chang H. Mental health and psychosocial problems of medical health workers during the COVID-19 epidemic in China. Psychotherapy and Psychosomatics. 2020;89(4):242–50. doi:10.1159/000507639

* Zhang Y, Ma ZF. Impact of the COVID-19 pandemic on mental health and quality of life among local residents in Liaoning Province, China: A cross-sectional study. International Journal of Environmental Research and Public Health. 2020;17(7):2381. doi:10.3390/ijerph17072381

* Zhao Y, An Y, Tan X, Li X. Mental health and its influencing factors among self-isolating ordinary citizens during the beginning epidemic of COVID-19. Journal of Loss and Trauma. 2020;25(6-7):580–93. doi: 10.1080/15325024.2020.1761592

* Zhu J, Sun L, Zhang L, Wang H, Fan A, Yang B, Li W, Xiao S. Prevalence and Influencing Factors of Anxiety and Depression Symptoms in the First-Line Medical Staff Fighting Against COVID-19 in Gansu. Frontiers in Psychiatry. 2020;11. doi:10.3389/fpsyt.2020.00386

