## Supplement A & B for "Mental Disorder Prevalence Among Populations Impacted by Coronavirus Pandemics: A Multilevel Meta-Analytic Study of COVID-19, MERS & SARS"

**Mental Disorder Measures**

| **Measure** | **Number of Studies in which Included** |
| --- | --- |
| Beck Anxiety Inventory (BAI) | 1 |
| Beck Depression Inventory (BDI) | 3 |
| Brief Symptoms Rating Scale (BPRS) | 1 |
| Center for Epidemiologic Studies - Depression (CES-D) | 5 |
| Chinese Health Questionnaire (CHQ) | 3 |
| Clinician Rating | 3 |
| Depression & Anxiety Stress Scale (DASS) | 5 |
| Davidson Trauma Scale (DTS) | 2 |
| Generalized Anxiety Disorder-7 (GAD-7) | 9 |
| General Health Questionnaire (GHQ) | 11 |
| Hospital Anxiety & Depression Scale (HADS) | 5 |
| Hamilton Anxiety Scale (HAS) | 1 |
| Hamilton Depression Scale (HDS) | 1 |
| Impact of Events Scale (IES) | 28 |
| Kessler-10 | 2 |
| MINI Neuropsychiatric Inventory | 2 |
| PTSD Checklist (PCL) | 8 |
| Psychological Well-being Questionnaire | 1 |
| Patient Health Questionnaire (PHQ) | 7 |
| Structured Clinical Interview for DSM (SCID) | 4 |
| Symptom Checklist-90 (SCL-90) | 1 |
| State and Trait Anxiety Inventory (STAI) | 2 |
| Zung's Self-rating Anxiety Scale | 3 |
| Zung's Self-rating Depression scale | 3 |

**Supplement B**

**Bias Coding**

The Systematic Assessment of Quality in Observation Research (SAQOR) tool provides criteria keyed to domains that are rated to obtain a multi-faceted quality assessment of individual studies. Domains include Sample, Control/Comparison Group, Quality of Exposure/Outcome, Follow-up (for longitudinal studies), Distorting Influences (i.e., key confounds), and Reporting of Data. We adhered as closely as possible to existing guidelines (Ross et al., 2011), while modifying guidelines to improve the applicability of the system for our purposes.

Sample domain

1. The sample was considered representative of the population from which it was drawn if: (a) the study reported evidence of representativeness obtained through analyses comparing the study sample to a normative sample; or (b) the study sample was recruited through consecutive or random sampling with at least 60% of eligible participants consenting to participate.
2. We did not code the criterion ‘The sample size is appropriate to identify statistical differences between groups for primary outcomes’, as group differences were not the focus of our analyses.
3. The criterion ‘Entry criteria stated and justified’ was coded ‘yes’ if inclusion/exclusion criteria were clearly stated and self-evident (e.g., all doctors/nurses working at a hospital impacted by SARS were recruited to examine prevalence of PTSD among healthcare providers).
4. A summary score of ‘adequate’ was provided if three of four criteria were coded ‘yes’, and ‘unclear’ if three of four were coded ‘unclear’.

Control/Comparison Group domain

5. A control/comparison group is not essential to reporting of point-prevalence data (as opposed to risk ratios). Thus, we did not code this domain, and instead coded presence of a control/comparison group as a key confounder under ‘Distorting Influences.’

Quality of Exposure/Outcome domain

6. Criterion ‘Adequate assessment of exposure’ was coded ‘unclear’ for the quarantined population when quarantine was self-reported and not objectively measured.
7. Criterion ‘Adequate measure of outcomes’ was coded in regard to mental disorder prevalence and not in regard to any other reported data (e.g., long-term functioning, physical outcomes).

Distorting Influences domain

8. We coded two distorting influences/key confounds: (1) prior mental disorders of sample, and (2) control/comparison group that serves as adequate control (e.g., non-quarantined control group when investigating mental disorders among quarantined sample).

Reporting of Data domain

9. Coding of this domain was keyed to mental disorder prevalence data only, and not any other analyses/data reported (e.g., group comparisons, regression analyses).
10. Criterion ‘Explanation for missing data is given’ was coded ‘no’ when (1) no explanation for missing data was reported, and (2) reported prevalence estimate differed from manually calculated estimate (sample *N* positive/total sample) - thus indicating participants were missing from the denominator of the reported prevalence estimate.
11. Criterion ‘Explanation for missing data is given’ was coded as ‘unclear’ when prevalence estimate was provided without number of participants positive for mental disorder. In these cases, it could not be confirmed whether provided prevalence estimate differed from manually calculated estimate, thus indicating missing data.
12. Criterion ‘Data are clearly and accurately presented including CI where appropriate’ was coded ‘yes’ if both *N* positive and prevalence (%) were reported.

Summary Scoring

13. Consistent with Ross and colleagues (2011), each domain was scored as ‘adequate’, ‘inadequate’ or ‘unclear’. Consistent with GRADE criteria as applied to observational studies (GRADE working group, 2004), all studies were initially considered ‘low’ quality evidence. Studies were moved into higher or lower quality evidence categories depending on the number of domains scored as adequate/inadequate/unclear.
    a. Studies were coded as providing moderate quality evidence when four or more domains were coded ‘adequate.’
    b. Studies were coded as providing very low quality when three or more domains were coded as ‘inadequate’ and/or ‘unclear.’
